# Black flies and Onchocerciasis: Knowledge, attitude and practices among inhabitants of Alabameta, Osun State, Southwestern, Nigeria

**DOI:** 10.1101/2020.11.13.20230995

**Authors:** Busari Lateef Oluwatoyin, O.O Ojurongbe, M.A Adeleke, O.A Surakat

## Abstract

**Background and Objectives:** This study reports knowledge of residents of Alabameta community, Osun State, Nigeria on the bioecology and socio-economic burden of black flies and onchocerciasis.

**Methods:** Using structured questionnaires and Focus Group Discussion (FGD), a total of 150 community respondent participated in the study.

**Results:** The knowledge of the residents on the existence of black flies in the community was significant (p<0.05) as all the 150 respondents confirmed the presence of black flies with the local name ‘Amukuru’. However, their lack of knowledge of the flies breeding site (104) (69%), prevention (134) (89%), cause (132) (88%), and treatment (133) (89%) of onchocerciasis was profound. Majority 147(98%) of the respondents reported that flies bite more in the wet season as against dry season 3(2%). The flies have a higher affinity (124) (82%) for biting the leg than any other part of the body. A larger percentage (89%) of the respondents are unaware of any medication for the treatment of onchocerciasis while 11% are aware. There had been no sensitization on onchocerciasis according to 89% of the respondents.

**Conclusion:** Due to lack of resident’s knowledge on black flies bioecology which may continuously expose them to the bite of the flies and ultimately infection, it is paramount that the Osun State government map out strategizes through orientation and drug administration with a view to onchocerciasis prevention and eradication.

## Introduction

Among the filarial nematodes, the public health significance of *Onchocerca volvulus* cannot be overemphasized being the causative organism of the dreadful and debilitating disease onchocerciasis. Human onchocerciasis (river blindness) causes blindness and severe dermatitis in Africa and Latin America (WHO, 1976) and is the second leading cause of blindness (Dent and Kazura, 2011). Onchocerciasis is transmitted by members of *Simulium damnosum* complex through their bite while taking a blood meal (Yameogo *et al*., 1999). Nine sibling species of *Simulium damnosum* complex have been taxonomically identified and documented in West Africa. The species include *Simulium sirbanum, S. damnosum sensu stricto, Simulium dieguerense, Simulium sanctipauli, Simulium soubrense, Simulium squamosum, Simulium yahense, Simulium leonense, Simulium konkorense* (WHO, 1994). The first three species are known as Savannah flies which transmit Savannah strain of *O. volvulus* while the rest belong to the forest group and transmit the Forest strain of the parasite which causes more of skin diseases than blinding disease (Mafuyai *et al*., 1996; Ibeh *et al*., 2006). The congregation of adult worms and subsequent fertilization to produce microfilariae in the subcutaneous tissues has been known to elicit nodules (Opara *et al*., 2005a).

It is known to be endemic in many tropical countries and over 18 million people are infected worldwide and 120 million people are at the risk of the disease (WHO, 1995). Onchocerciasis is most common in Africa while Nigeria probably has the highest burden of the disease (Oyibo and Fagbenro, 2003). Furthermore, Southwestern Nigeria (according to APOC rapid epidemiological mapping in 2008) was identified as one of the onchocerciasis endemic regions of the country. Earlier epidemiological studies showed that Alabameta is highly endemic with onchocerciasis with low community microfilarial load (Ojurongbe *et al*., 2015).

The present study therefore provides information on knowledge, attitude and practice of residents of Alabameta, Osun State, Nigeria on bioecology and socio-economic burden of black flies with a view to understanding its public health implication and in planning effective control strategies in the study area and Osun State, Nigeria at large.

## Materials and Methods

### Study Area

The Community is located in Ife South Local Government of Osun State, Southwest, Nigeria with an area of 730 kilometer square and a population of 135,338(2006 census). Owena River transverses the community and extends to Ondo State. The river usually produces rapids during the wet season and serves as a conducive breeding site for *S. damnosum*. However, during the dry season, the volume of water is usually drastically reduced to the extent that the rocks in the river are visible but does not become completely dry at all. Water from the river is used by residents to serve their domestic needs such as bathing and washing.

Alabameta lacks the necessary social amenities such as electricity, pipe borne water, hospitals, good roads, mobile phone and internet network and recreation centers expected in a human habitat. The major occupation of residents is farming.

### Ethical Clearance

A written consent was sought and obtained from the Osun State Ministry of Health.

### Questionnaire Administration and Focus Group Discussion (FGD)

150 Structured questionnaires were administered to community members that are fifteen years and above to assess their knowledge on the bioecology and socioeconomic burden of the black fly. The questions captured the place of bite, season/month with the highest biting incidence and time, breeding site, effects of bite, body part mostly bitten and ways adopted to preventing fly bite. The questionnaires were complemented with Focus Group Discussion (FGD) of seven participants which included old men, old women and youths. The questions for the focus group discussion were unstructured and targeted at seeking information about the resident’s general knowledge on the bioecology, attitudes and practice against black flies.

### Data Analysis

The data from the study was subjected to T-test and Chi-square to determine the significant difference in dynamics and transmission potential of black flies during the period of the study. All analysis was performed using SPSS version 17.

## Results

A total of 150 respondents were interviewed. Respondents within the age bracket 31-50 (69) (46%) were higher than those within the age brackets 15-30 (54) (36%) and 51 above (27) (18%). There were more male respondents (76) (51%) than female respondents (74) (49%). About 51% (76) of the respondents were without formal education while 6% were educated up to tertiary level. Farming was the major occupation (48%), followed by trading (26%) and fishing (11%) while students made up the rest of the respondents (8%) (Table 1).

**Table 1:** Demographic data of respondents of the community

| Characteristic | Age group | No. of respondent | Percentage % |
| --- | --- | --- | --- |
| Age | 15-30 | 54 | 36 |
|  | 31-50 | 69 | 46 |
|  | 51 above | 27 | 18 |
| Gender | Male | 76 | 51 |
|  | Female | 74 | 49 |
| Educational Level | Primary | 31 | 20 |
|  | Secondary | 34 | 23 |
|  | Tertiary | 9 | 6 |
|  | None | 76 | 51 |
| Occupation | Farming | 73 | 48 |
|  | Fishing | 16 | 11 |
|  | Trading | 39 | 26 |
|  | Student | 12 | 8 |
|  | Others | 10 | 7 |

Table 2 summarizes the knowledge and attitude of respondents on bioecology of *S. damnosum* complex. All the respondents 150 (100%) affirmed that they know black flies when probed of the presence of the fly in the community with its local or common name ‘Amukuru’. They unanimously established the fact that the flies bite seriously in the community. A larger percentage (73%) of the respondents established that the flies bite mostly at the riverside (Owena River), while 27% claimed they bite mostly at the farm. Information from the Focus Group Discussion (FGD) corroborated these observations. The flies were believed to be biting any part of the exposed body but with affinity for the leg (124) (82%) than any other part of the bodies. This is also affirmed during the FGD. The major effect of the fly’s bite was attributed to itching by the respondents (80%) while 28(19%) respondents claimed it causes swelling.

**Table 2:** Knowledge and attitude of respondents on bioecology of *Simulium damnosum* complex

| Parameter | Response | No. of Respondent | Percentage |
| --- | --- | --- | --- |
| Have you heard of black flies? | Yes | 150 | 100 |
|  | No | 0 | 0 |
| Do they bite in your community? | Yes | 150 | 100 |
|  | No | 0 | 0 |
| Where do they bite in the community? | Farm | 38 | 25 |
|  | Riverside | 109 | 73 |
|  | House | 2 | 1 |
|  | Others | 1 | 1 |
| Which part of the body do they often bite? | Head | 9 | 6 |
|  | Neck | 13 | 9 |
|  | Leg | 124 | 82 |
|  | Others | 4 | 3 |
| What are the effects of their bite? | Itching | 121 | 80 |
|  | Swelling | 28 | 19 |
|  | Others | 1 | 1 |
| Where do they breed? | River | 46 | 31 |
|  | Tree | 104 | 69 |
|  | Others | 0 | 0 |
| Which season do they bite most? | Wet | 147 | 98 |
|  | Dry | 3 | 2 |

About 69% of the respondents claimed that black flies breed in tree holes. Respondents during FGD also ascribed the breeding of black flies to tree holes such as ‘Iroko kekere’ and ‘Igi araba”. Most of the respondents confirmed that black flies bite more in the wet season 147(98%) as against dry season 3(2%).

Table 3 shows that 88% of the respondents were ignorant of black flies as vectors of onchocerciasis. The level of ignorance was also evident among participants in the Focus Group Discussion (FGD). 36% believed onchocerciasis is caused as a result of mosquito bite, 23% attributed it to witchcraft, 11% claimed it’s the consequence of eating kola nut, 3% see it as being hereditary while 27% have no idea of its cause.

**Table 3:**
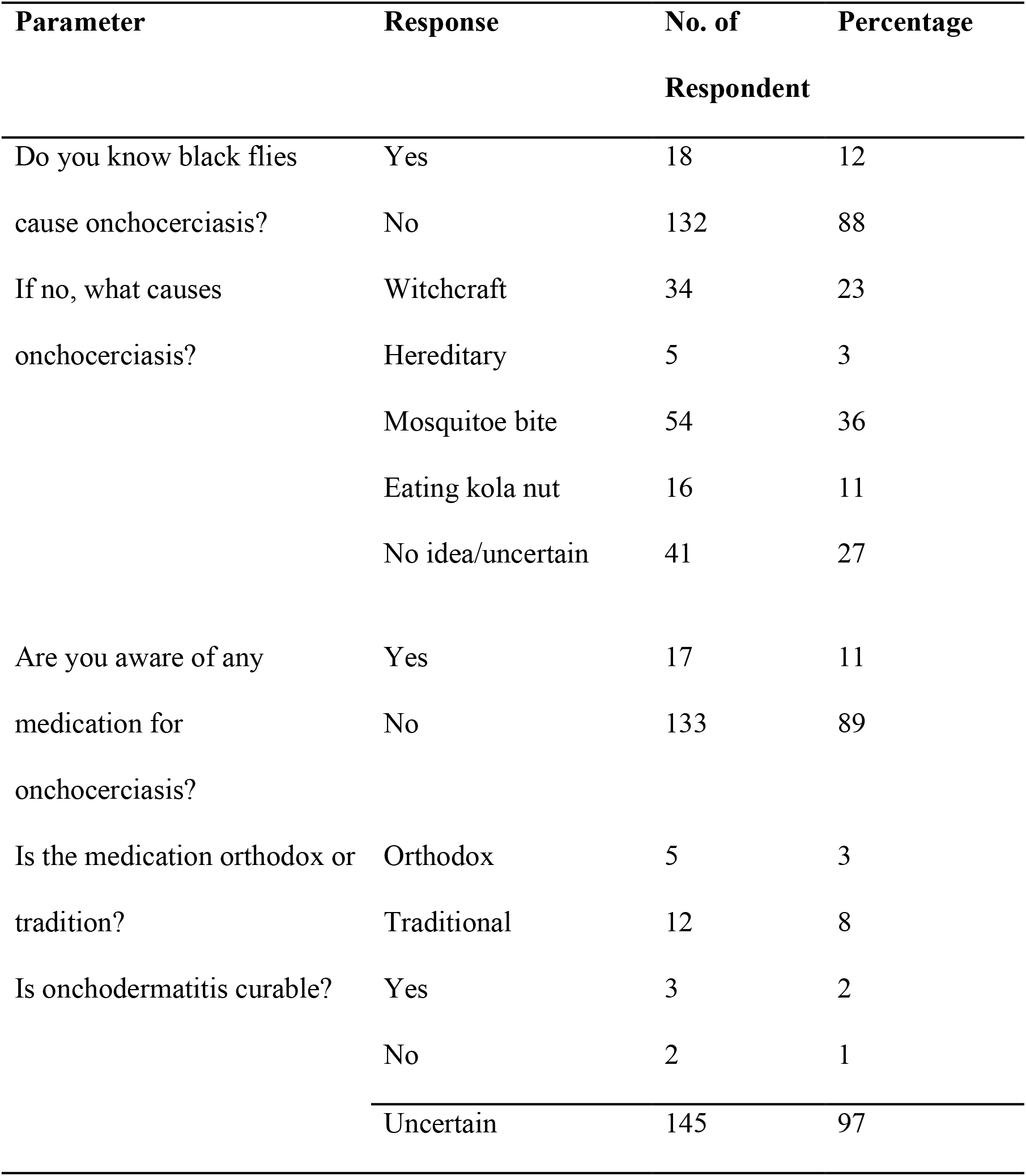
Knowledge of respondents on onchocerciasis, causes and treatment

A larger percentage (89%) of the respondents is unaware of any medication for the treatment of onchocerciasis while 11% are aware. However, few respondents claimed the use of orthodox medicine while 8% claimed it is traditional.

Table 4 summarizes the knowledge of respondents on methods of black flies bites prevention. There had hardly been any sensitization on onchocerciasis according to 89% of the respondents and later confirmed during the FGD. However, 91 (60%) of respondents prevent fly bite by clothing their bodies (such as wearing socks, long garments and sweater) while 52 (35%) rub ointment such as palm oil or mosquito repellant creams. Biting is usually intense in the morning according to 83% of the respondents, followed by evening (24) (16%) while fly bite in the afternoon is the least (1) (1%). After fly bites, majority 48% do nothing apart from killing the fly, 10% rub ointment, 7% go to the hospital while 4% take drugs (particularly antimalarial drugs).

**Table 4:** Knowledge of respondents on methods of prevention of black flies bite

| Parameter | Response | No. of Respondent | Percentage |
| --- | --- | --- | --- |
| Has there been any sensitization on onchocerciasis? | Yes | 16 | 11 |
|  | No | 134 | 89 |
| How do you prevent the flies from biting? | Clothing | 91 | 60 |
|  | Rubbing of ointment | 52 | 35 |
|  | Others | 7 | 5 |
| What time of the day is biting intense? | Morning | 125 | 83 |
|  | Afternoon | 1 | 1 |
|  | Evening | 24 | 16 |
| What do you do after fly bite? | Take drugs | 6 | 4 |
|  | Go to hospital | 11 | 7 |
|  | Rub ointment | 15 | 10 |
|  | Nothing | 71 | 48 |
|  | Others | 47 | 31 |

## Discussion

The result on perception of the community residents on bioecology of black flies showed that they are conversant with the black fly bites. A larger population of the residents affirmed that the flies bite mostly along the river side corroborating the scientific findings that people working close to the rivers are at the high risk of *Simulium* biting nuisance and onchocerciasis (Akogun and Onwuliri, 1991; Abdullahi and Oyeyi, 2003). The reason for the impressive knowledge of the daily and seasonal distribution of the flies could be attributed to the long time presence of *Simulium* in the area. However, the river serves as a major source of water for domestic and occupational demand. Therefore reducing man-fly contact by refraining from such high biting areas of *Simulium* is paramount in controlling onchocerciasis.

The knowledge of the residents revealed that the flies bite mostly in the wet season than in the dry season which is in consonance with findings by Adeleke *et al*., (2010b). This indicates that there is a higher risk of exposure to onchocerciasis in the wet season than in the dry season due to increase in adult fly population during the wet season. Thus, disease control is best carried out in the dry season since it gives a better chance for the destruction of the breeding site which invariably leads to the eradication of the flies. The poor knowledge of the endemic communities on the ecology of *S. damnosum s*.*l* had also been documented in many parts of Nigeria (Anosike and Onwuluri, 1995; Ukpai and Ezeji, 2003; Dozie *et al*., 2004).

Also, the poor knowledge of the residents on the breeding sites of *S. damnosum s*.*l* could increase their risk to onchocerciasis as majority of the residents do not have accurate knowledge of their breeding sites. Majority of residents attributed their breeding site to a tree “Iroko kekere” or “Igi araba” and as such do not really pay attention to fly bite.

Itching according to a larger population of the resident is one of the effects of fly bite which could be painful, and at times accompanied with blood from the site of bite while trying to kill the fly. It is one of the recognized early manifestations of onchocerciasis and could in many severe conditions, lead to skin lesions and disfigurement in affected individuals (Adewale *et al*., 1999; Ukpai and Ezeji, 2003).

Furthermore, 88% of the residents do not have knowledge regarding the cause of onchocerciasis as many claimed that fly bite causes malaria. This is as a result of lack of sensitization and formal education and could lead to the persistence of the onchocerciasis in the community. Public health enlightenment on black fly and onchocerciasis is therefore paramount for community members in a view to preventing the disease. The cooperation of the inhabitants in endemic villages is needed besides the currently employed mass chemotherapy required to control human onchocerciasis in Nigeria.

Since the residents lack knowledge of the cause of onchocerciasis, it is not unlikely their ignorance of the treatment for the disease as shown in the result. There is therefore the need for urgent sensitization on the treatment of the disease through the distribution of ivermectin to residents who possibly show the disease symptoms. There is also the need for them to be acquainted not only with the available medications and modern tools for controlling human onchocerciasis but with an acceptable way of knowing the causative agent, mode of transmission and preventive measures. Visual demonstrations of parasite development in *Simulium* flies, transmission through their bites and identification of adult parasites in excised nodules may prove to be an effective tool in a health education programme.

This is paramount due to their lack of seeking medical attention when bitten by the flies possibly due to the unavailability of hospital in the environment or their use of antimalarial drugs since they believe that black flies cause malaria.

Although, many prevent the flies from biting by wearing clothings that cover their legs properly (p=0.088; p>0.05), they usually complain of being uncomfortable with such clothings particularly while working in the farm which more often than none makes them wear shorts and get bitten by the flies thus exposing them to the disease. This is coupled with the fact that they claim the fly bite do not prevent them from work or going out for their daily activities (p=0.304; p>0.05).

## Data Availability

All relevant data are included in the manuscript

## Acknowledgement

I wish to express my profound gratitude to my esteemed teachers and mentors Dr. Ojurongbe O.O and Dr. Adeleke M.A for their distinctive tutelage. Also, is my mum and family. I appreciate all those who have contributed in one way or the other to the accomplishment of this work.

